# The Health and Economic Impact of the Adult-specific 21-valent (aPCV21) Pneumococcal Conjugate Vaccine vs PCV20 on Invasive Pneumococcal Disease Among Adults Aged 65 years in the United Kingdom: A Modeling Study

**DOI:** 10.64898/2026.10.03.26364601

**Authors:** Peter P. Mueller, Muloongo Simuzingili, Ian R. Matthews, Zinan Yi, Kwame Owusu-Edusei

## Abstract

**Introduction:** This study compared the health and economic impact of the 21-valent pneumococcal conjugate vaccine (aPCV21) vs the 20-valent pneumococcal conjugate vaccine (PCV20) on invasive pneumococcal disease (IPD) and post-meningitis sequelae (PMS) among adults aged 65 years and older in the United Kingdom (UK).

**Methods:** A published Markov model was used to simulate a single-cohort of vaccine-naïve adults who became eligible for age-based vaccination at age 65 years, with uptake occurring over time. The cohort was followed until a maximum age of 100 years. Model inputs were derived from publicly available national health data and published reports. Outcomes were discounted at 3.5% annually, and costs were adjusted to the 2024 Great Britain Pound (£). Additional analyses explored the impact of changing the age-based recommendation from 65 years to 75 years. Scenarios involving instantaneous vaccine uptake rates were included as part of the sensitivity analyses. A deterministic sensitivity analysis was performed on the incremental direct medical costs between vaccinating with aPCV21 vs PCV20 under the age-based recommendation of 65 years.

**Results:** Using an age-based recommendation of 65 years, aPCV21 led to fewer cases, deaths, and costs than PCV20. Compared with PCV20, aPCV21 averted an additional 138 IPD cases, 1 PMS case, and 22 IPD deaths, saving £1.2 million in direct medical costs. Raising the age-based recommendation from 65 to 75 years and using aPCV21 (or PCV20, respectively), resulted in 120 (92) additional IPD cases, 2 (1) additional PMS case(s), 19 (14) additional IPD death(s), and an extra £1.1 (£0.8) million in direct medical costs. The deterministic sensitivity analysis showed that aPCV21 reduced direct medical costs across all parameter ranges.

**Conclusions:** Compared with PCV20, aPCV21 prevented more IPD cases, IPD deaths, and direct medical costs among older adults in the UK. Changing the age-based recommendation from age 65 to 75 resulted in worse health and economic outcomes.

## Introduction

*Streptococcus pneumoniae* is a gram-positive bacteria transmitted through respiratory secretions, which causes pneumococcal disease (PD), a potentially severe or fatal condition that includes non-bacteremic pneumococcal pneumonia (NBPP) and invasive pneumococcal disease (IPD).^1^ *S. pneumoniae* capsular polysaccharides are key determinants of pathogenicity and host immune response, and thus have been used to classify pneumococci by serotypes. More than 100 serotypes have been identified to date.^1^ The global burden of PD equates to 197 million cases and over 118,000 deaths annually,^2^ with children aged <5 years, immunocompromised individuals, and older adults being the most affected population groups.^1^ In the United Kingdom (UK), IPD incidence is 18.0 per 100,000 among adults aged 65 to 70 years and 37.5 per 100,000 among adults aged ≥80 years.^3^ Vaccination is the cornerstone of PD prevention.^4^ Between 2003 and 2026, the UK recommended routine vaccination with the 23-valent pneumococcal polysaccharide vaccine (PPSV23) for adults aged ≥65 years.^5,6^ PPSV23 is comprised of purified polysaccharide antigens of 23 bacterial serotypes, while pneumococcal conjugate vaccines (PCVs) use a nontoxic variant of diphtheria toxin to anchor a varying number of *S. pneumoniae* serotype saccharides.^4^ However, unlike PCVs, PPSV23 does not prevent carriage, is only moderately effective, and has rapid waning of protection,^7,8^ limiting its preventive value among older comorbid adults.^9^ For this reason, PCV20 replaced PPSV23 for the adult national immunization program in the UK in early 2026.^1,10^

A high burden of non-vaccine type-serotype pneumonia continues to affect older adults,^11,12^ highlighting the need for a PCV specifically developed for the adult population.^5^ Addressing this issue, Merck & Co., Inc., Rahway, NJ, USA has introduced aPCV21, a 21-valent PCV designed specifically for adults, which includes pneumococcal serotypes 3, 6A, 7F, 8, 9N, 10A, 11A, 12F, 15B, 17F, 19A, 20A, 22F, and 33F, as well as 8 unique serotypes not currently contained in any licensed vaccine: 15A, 15C [from deOAc-15B], 16F, 23A, 23B, 24F, 31 and 35B.^13^ These 8 unique serotypes account for over 19% of IPD cases among UK adults aged ≥65 years and have been identified in severe or fatal IPD outcomes among older adults in the UK.^12^ Overall, aPCV21 serotypes account for approximately 84% of IPD cases among adults aged ≥65 years, whereas PCV20 serotypes account for only 60% of such cases.^14^

In this context, epidemiological modeling based on real-world data can help elucidate the health impacts of using aPCV21 or PCV20 to vaccinate older adults in the UK, providing essential data to inform the UK’s national vaccination program. Other key information needed in this area includes the cost implications of vaccine types, as cost-effectiveness has been stipulated as a criterion for health interventions within the UK’s publicly funded medical system.^15^ It has been suggested that the UK’s current age-based recommendation for pneumococcal vaccination (set at age 65) be shifted to age 75.^16^ However, the clinical and cost implications of such a change remain unknown. The primary argument for shifting the age-based recommendation to age 75 is to increase protection at the age when incidence is highest (PD risk increases with age in adults^3^). But the trade-off with this approach would be the large number of unprotected adults aged 65-74 years. To address a need for more information on adult pneumococcal vaccination strategies in the UK, the current study estimated the health and economic outcomes of aPCV21 and PCV20 across two age-based vaccination strategies in older adults.

## Methods

### Model overview

Using a previously published Markov model,^17^ we evaluated the health and economic impacts of vaccination with aPCV21 compared with the current program of PCV20 among older adults in the UK. In all analyses, the model followed a single-cohort of vaccine-naïve adults starting from the age of 65 years through a maximum age of 100 years, using an initial population size of 689,198 people. As they aged, individuals could transition between the following states: no IPD, IPD, post-meningitis sequelae (PMS), and death (**Supplemental Figure S1**). The cohort was stratified into 3 mutually exclusive risk groups: low-risk, at-risk, and high-risk. Individuals began in the no IPD state. A proportion of IPD cases were assumed to lead to meningitis (**Table 1**). In addition, a proportion of meningitis cases were assumed to lead to PMS, which was assumed to last for the rest of individual’s lifetime. Non-meningitis IPD and meningitis IPD that did not result in PMS were set to resolve within 1 year (the length of 1 cycle). Individuals exited the model at death, based on either the age-specific background mortality rate or the case fatality rate for the IPD state. A half-cycle correction was applied to cumulative cost outcomes.

**Table 1.**
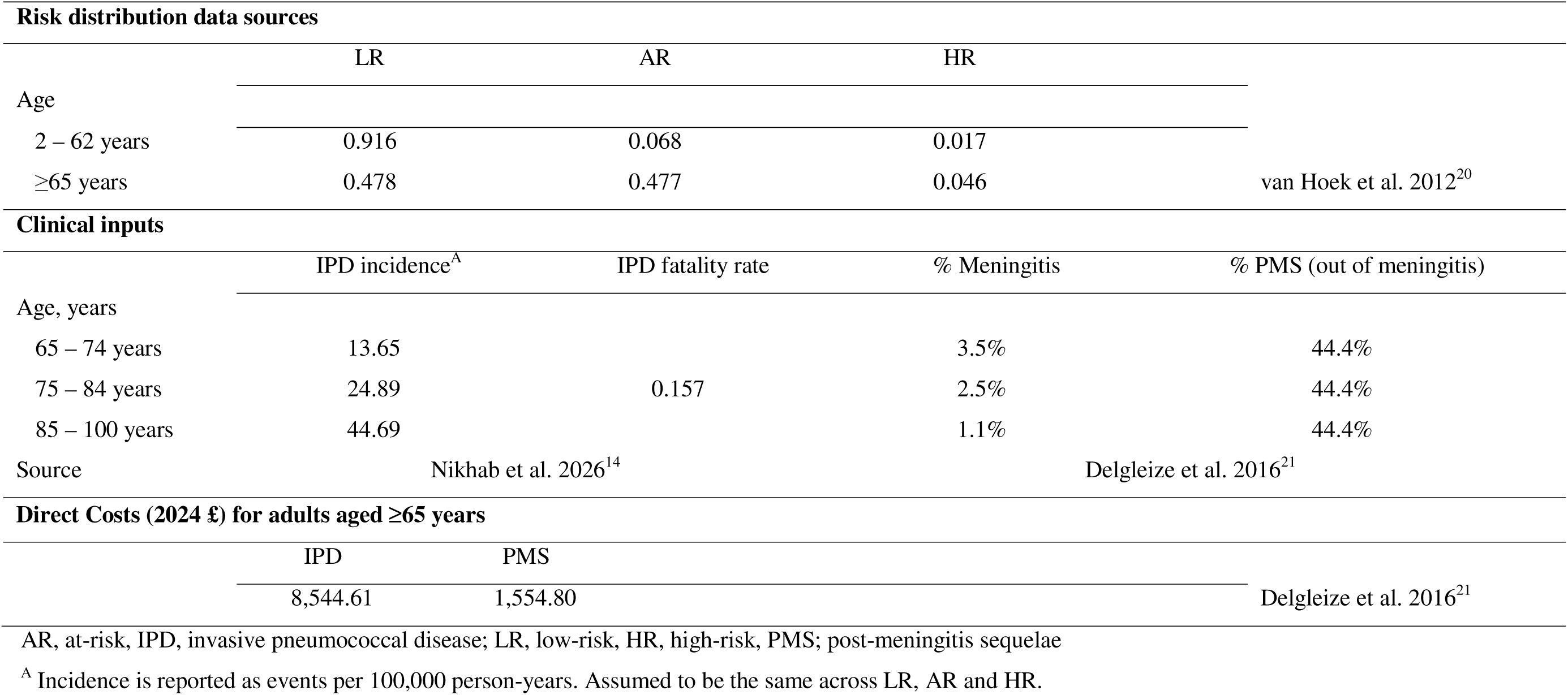
Clinical and economic model inputs.

### Model input and data sources

Population size^18^ and background mortality rates^19^ for each age were based on UK census data (**Supplemental Table S1**). Age-specific incidence rates of IPD per 100,000 person-years were derived from the case counts reported in a UK study by Nikhab et al. 2026.^14^ Details on risk condition distributions and progression within the model are outlined in the supplement. The risk-distributions were taken from the findings of a real-world study of hospitalized IPD patients in the UK.^20^ Previously published cost-effectiveness data on pneumococcal vaccination in the UK^21^ were used to determine the direct medical costs per case of IPD and PMS. Health and economic outcomes were discounted at an annual rate of 3.5%.^22^ All costs were inflation adjusted to the 2024 Great Britain Pound (GBP) using the consumer price index from the Office for National Statistics.^23^

The model allowed for vaccination at any year in the lifetime horizon of the single-cohort of adults aged 65 years, but revaccinations were not allowed. Vaccine uptake rates refer to the rate at which people were vaccinated within a given annual cycle in the model. Age-specific vaccine uptake rates were based on data in the 2024-2025 Pneumococcal Polysaccharide Vaccine Coverage Report^24^ and assumed to be equal for aPCV21 and PCV20 (**Supplemental Table S2**). Age-specific vaccine uptake rates are referred to as “real-world uptake rates”. In the base case, 15.4% of the cohort were vaccinated at age 65, a further 12.4% were first vaccinated at age 66, and so on (following trends in **Supplemental Table S2**).

The distribution of pneumococcal serotypes among IPD cases was derived from data reported in Nikhab et al. 2026^14^ (**Supplemental Table S3**). Serotype-specific vaccine efficacy was assumed to be the same for aPCV21 and PCV20 but to be lower in high-risk individuals than in low-risk or at-risk individuals (**Supplemental Table S4**).^25^ Vaccine efficacy against serotype 3 was assumed to be significantly lower than the other covered serotypes. The duration of protection was assumed to be 15 years for both aPCV21 and PCV20, with full efficacy for the first 5 years followed by a linear decrease from year 6 onwards, reaching 0% efficacy by year 15.^7^

### Model outcomes

The model estimated the health and economic outcomes associated with IPD under vaccination with either aPCV21 or PCV20. Health outcomes were IPD cases, PMS cases, and IPD deaths. Economic outcomes were direct medical costs calculated by applying cost inputs to each case of IPD or PMS. The base case analysis estimated the impact of vaccinating with either aPCV21 or PCV20 under the current age-based recommendation of 65 years using the real-world uptake rates.

### Scenario analysis: Health and economic outcomes of beginning vaccination at 75 years compared with 65 years

A scenario analysis was conducted to evaluate the impact on projected model outcomes of changing the age-based vaccination recommendation from 65 years to 75 years for both aPCV21 and PCV20. The analysis followed a single-cohort of vaccine-naïve adults from the age of 65 years through a maximum age of 100 years with the age-based vaccination recommendation starting at age 75. In the scenario analysis, only risk-based vaccination occurred between the ages of 65 and 74. An annual uptake rate of 22.9% was assumed for those “newly registered as ‘at-risk’” across ages 65-74 (where ‘at-risk’ in the PPV Coverage Report corresponds to the combined at-risk and high-risk subgroups of our model).^24^ Of those who did not receive risk-based vaccination between the ages of 65 and 74, 15.4% got their first vaccine at age 75, a further 12.4% received their first vaccination at age 76, and so on, following trends described in **Supplemental Table S2.** Clinical and cost outcomes for the scenario analysis were compared with those of the base case analysis in which age-based vaccination started from age 65.

### Scenario analysis: Impact of vaccine uptake rate assumptions

An alternative set of vaccine uptake rates were also explored, in which vaccine administrations were focused on a single-age. Using assumptions derived from real-world data^24^ an instantaneous uptake rate of 73.2% was applied at either age 65 or 75, depending on the age-based recommendation scenario. This alternative set of uptake rates (which we refer to as “instantaneous uptake rates”) was intended to measure the impact of changing the age-based recommendation under strict application of the policy instead of how it is currently being realized.

### Scenario analysis: Maximal vaccine impact and optimal vaccination age

As an additional exercise, we conducted a vaccine impact analysis like that performed in a modeling study by Thindwa et al. 2023.^16^ To determine the optimal vaccination age, Thindwa et al. estimated the total IPD cases averted as a function of vaccination age. In a similar manner, we used our model to estimate total IPD cases averted as a function of vaccination age, with 5-year intervals, with and without the 3.5% discount rate for health outcomes. This analysis was performed for both aPCV21 and PCV20 under the instantaneous uptake rate assumption with 100% vaccine uptake. The number of averted cases was computed by comparing outcomes with a 0% vaccine uptake scenario (referred to as “No vaccine”).

### Sensitivity analysis

A one-way deterministic sensitivity analysis (DSA) was performed on the base case to determine which parameters had the most substantial impact on total incremental direct medical costs. Vaccine efficacy against IPD was varied around base case values for each vaccine type and risk level (**Supplemental Table S4**). Other model parameters were varied by ± 20% of base case values. Results were presented in a tornado diagram as incremental costs of aPCV21 vs PCV20.

## Results

### Health and economic outcomes of beginning vaccination at 65 years

A single-age cohort of 689,198 adults aged 65 years were simulated over a lifetime horizon using the current age-based recommendations of 65 years. Estimated health outcomes for aPCV21 and PCV20 are presented in **Table 2**, with the respective cost outcomes shown in **Figure 1**. Under the real-world uptake rate assumptions, considered to be the base case, vaccination with aPCV21 under an age-based recommendation of 65 years resulted in an estimated 1,785 IPD cases, 17 PMS cases, and 281 IPD deaths. Similarly, vaccination with PCV20 under an age-based recommendation of 65 years resulted in an estimated 1,923 IPD cases, 18 PMS cases, and 303 IPD deaths. Compared with PCV20, aPCV21 prevented an additional 138 IPD cases, 1 PMS case, and 22 IPD deaths. Under vaccination with aPCV21, direct medical costs were estimated to be £15.2 million from IPD and £258 thousand from PMS. Under vaccination with PCV20, direct medical costs were estimated to be £16.4 million from IPD and £279 thousand from PMS. Compared with PCV20, aPCV21 reduced direct medical costs by a total of £1.2 million.

**Figure 1.**
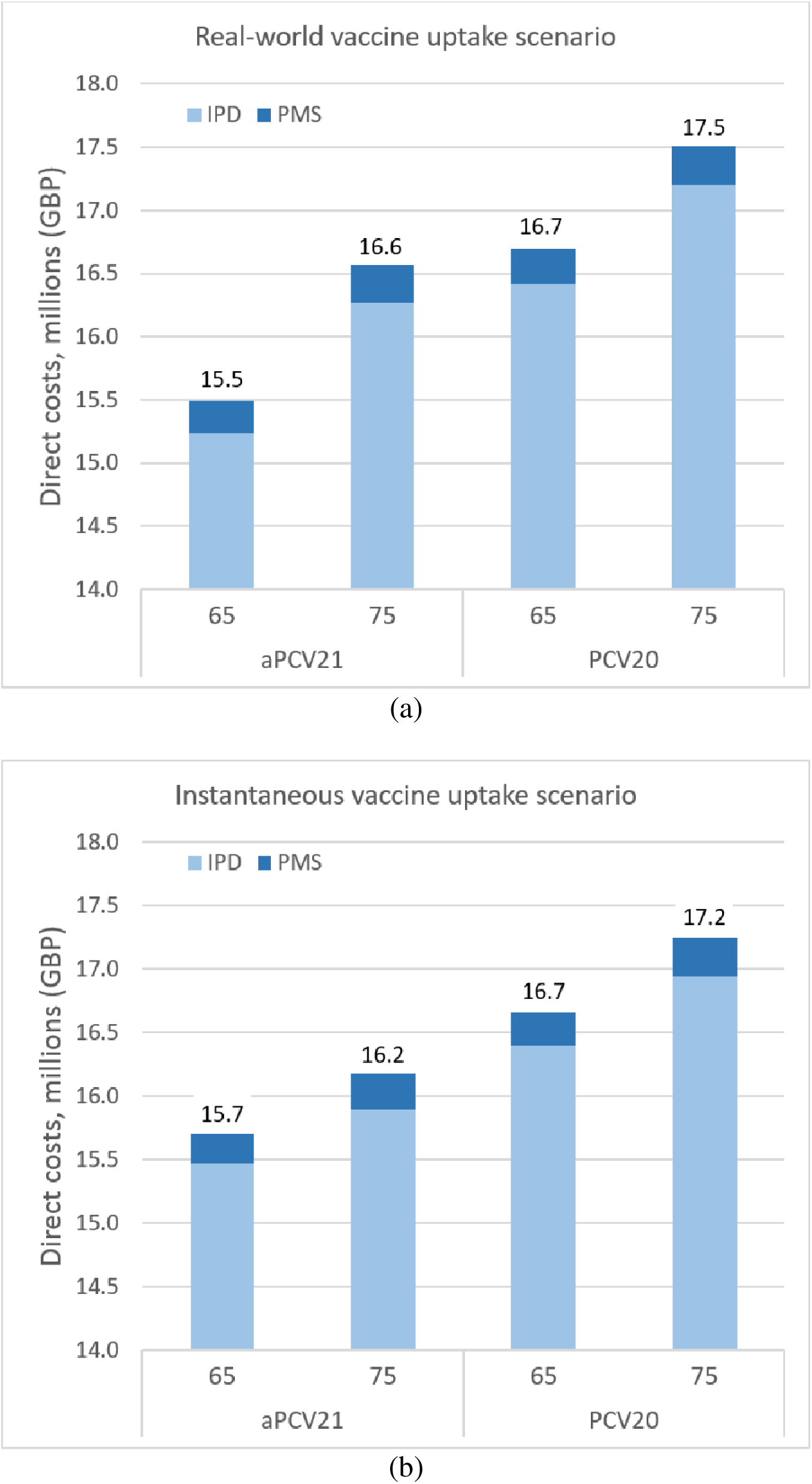
Predicted direct medical costs associated with treatment of invasive pneumococcal disease, by vaccine type and age-based recommendations. GBP, Great Britain pound; IPD, invasive pneumococcal disease; PMS, post-meningitis sequelae; aPCV21, adult-specific 21-valent pneumococcal conjugate vaccine; PCV20, 20-valent pneumococcal conjugate vaccine Bars represent lifetime direct medical costs estimated in the analysis of UK adults starting from age 65 years under the (a) real-world vaccine uptake rate assumption or the (b) instantaneous vaccine uptake rate (of 73.2%). Direct medical costs were adjusted to the 2024 GBP and were discounted at an annual rate of 3.5%. The categories of “65” and “75” correspond to the age-based recommendation options for pneumococcal vaccination in older adults. Data labels show total direct costs.

**Table 2.** Predicted health outcomes with vaccination starting at the age of 65 years, by pneumococcal vaccine type.

| Outcomes<br>(discounted) | Real-world uptake rates <sup>A</sup> |  |  | Instantaneous uptake rates <sup>B</sup> |  |  | No vaccine |
| --- | --- | --- | --- | --- | --- | --- | --- |
|  | aPCV21 | PCV20 | aPCV21 vs<br>PCV20 | aPCV21 | PCV20 | aPCV21 vs<br>PCV20 |  |
|  | Total number, n |  | Difference,<br>n | Total number, n |  | Difference,<br>n |  |
| IPD cases | 1,785 | 1,923 | -138 | 1,812 | 1,921 | -109 | 2,159 |
| PMS cases | 17 | 18 | -1 | 16 | 17 | -1 | 20 |
| IPD deaths | 281 | 303 | -22 | 285 | 302 | -17 | 340 |

IPD, invasive pneumococcal disease; PMS, post-meningitis sequelae; aPCV21, adult-specific 21-valent pneumococcal conjugate vaccine; PCV20, 20-valent pneumococcal conjugate vaccine
Table entries based on modeling of a single-cohort starting at the age of 65 years. All health outcomes were discounted at an annual rate of 3.5%.
<sup>A</sup> Real-world uptake rates use the values presented in Supplemental Table S2.
<sup>B</sup> Instantaneous uptake rates assume that 73.2% of vaccine-naïve 65-year-olds receive a vaccination at the start of the model. Further details are in the supplement.

### Scenario analysis results: Health and economic outcomes of beginning vaccination at 75 years compared with 65 years

The estimated health outcomes from shifting to an age-based recommendation of 75 years are presented in **Table 3**. Vaccination with aPCV21 under an age-based recommendation of 75 years resulted in an estimated 1,905 IPD cases, 19 PMS cases, and 300 IPD deaths. This equated to an increase of 120 IPD cases, 2 PMS cases, and 19 IPD deaths compared with aPCV21 vaccination under the current age-based recommendation of 65 years. Likewise, when the age-based recommendation was shifted from 65 to 75 years, vaccination with PCV20 resulted in an estimated 2,015 IPD cases, 19 PMS cases, and 317 IPD deaths. This equated to an increase of 92 IPD cases, 1 PMS case, and 14 IPD deaths compared with PCV20 vaccination under the current age-based recommendation.

**Table 3.** Scenario analysis: Predicted health outcomes with vaccination starting at the age of 75 years vs 65 years, using real-world uptake rates, by pneumococcal vaccine type.

| Outcomes<br>(discounted) | 75 age-based recommendation |  | 75 vs 65 |  |
| --- | --- | --- | --- | --- |
|  | aPCV21 | PCV20 | aPCV21 | PCV20 |
|  | Total number, n |  | Incremental difference, n |  |
| IPD cases | 1,905 | 2,015 | 120 | 92 |
| PMS cases | 19 | 19 | 2 | 1 |
| IPD deaths | 300 | 317 | 19 | 14 |

IPD, invasive pneumococcal disease; PMS, post-meningitis sequelae; aPCV21, adult-specific 21-valent pneumococcal conjugate vaccine; PCV20, 20-valent pneumococcal conjugate vaccine
Table entries based on modeling of a single-cohort starting at the age of 65 years. All health outcomes were discounted at an annual rate of 3.5%.
The incremental difference is calculated as the number of cases resulting from the 75 age-based recommendation, minus the cases resulting from the 65 age-based recommendation, each under the real-world uptake scenario.

The estimated direct medical costs under an age-based recommendation of 75 years are shown in **Figure 1**. Direct medical costs of vaccination with aPCV21 under an age-based recommendation of 75 years were estimated to be £16.3 million for IPD and £300 thousand for PMS. This equated to a total increase of £1.1 million compared with aPCV21 vaccination under the current age-based recommendation. If the age-based recommendation shifted from 65 to 75, direct medical costs under vaccination with PCV20 were estimated to be £17.2 million for IPD and £308 thousand for PMS. This equated to a total increase of £816 thousand compared with PCV20 vaccination under the current age-based recommendation.

### Scenario analysis results: Impact of vaccine uptake rate assumptions

In a single-cohort of adults aged 65 years, the estimated health outcomes from using either real-world or instantaneous uptake rate assumptions, under the age-based recommendation of 65 years, are shown in **Table 2**. Vaccination with aPCV21, under instantaneous uptake focused solely at 65 years of age, resulted in an estimated 1,812 IPD cases, 16 PMS cases, and 285 IPD deaths. Vaccination with PCV20, under instantaneous uptake focused solely at 65 years of age, resulted in an estimated 1,921 IPD cases, 17 PMS cases, and 302 IPD deaths. When compared to the health outcomes under the real-world uptake rate assumptions, as presented in the base case, the relative differences across the two vaccine uptake rate options reached a maximum of 5.9% (with aPCV21, PMS cases totaled 17 under real-world uptake rates vs 16 under instantaneous uptake rates).

Direct medical costs following vaccination with aPCV21, under instantaneous uptake focused solely at 65 years of age, were estimated to be £15.5 million from IPD and £238 thousand from PMS. Direct medical costs following vaccination with PCV20, under instantaneous uptake focused solely at 65 years of age, were estimated to be £16.4 million from IPD and £263 thousand from PMS. Under the age-based recommendation of 65 years and vaccination with aPCV21 (or PCV20, respectively), the impact of shifting from the real-world to instantaneous uptake rate assumptions yielded a 7.8% (5.7%) relative decrease in PMS medical costs and a 1.5% (0.1%) relative increase (decrease) in IPD medical costs (**Figure 1**).

In a single-cohort of adults aged 65 years, the estimated health outcomes from using instantaneous uptake rate assumptions, alongside the age-based recommendation of 75 years, are presented in **Table 4**. Vaccination with aPCV21, under instantaneous uptake focused solely at 75 years of age, resulted in an estimated 1,861 IPD cases, 18 PMS cases, and 293 IPD deaths. Vaccination with PCV20, under instantaneous uptake focused solely at 75 years of age, resulted in an estimated 1,985 IPD cases, 19 PMS cases, and 313 IPD deaths. When compared to the health outcomes under the real-world uptake rate assumptions alongside the age-based recommendation of 75 years (presented in **Table 3** and as part of the prior scenario analysis), the relative differences across the two vaccine uptake rate options reached a maximum of 5.6% (with aPCV21, PMS cases totaled 19 under real-world uptake rates vs 18 under instantaneous uptake rates).

**Table 4.**
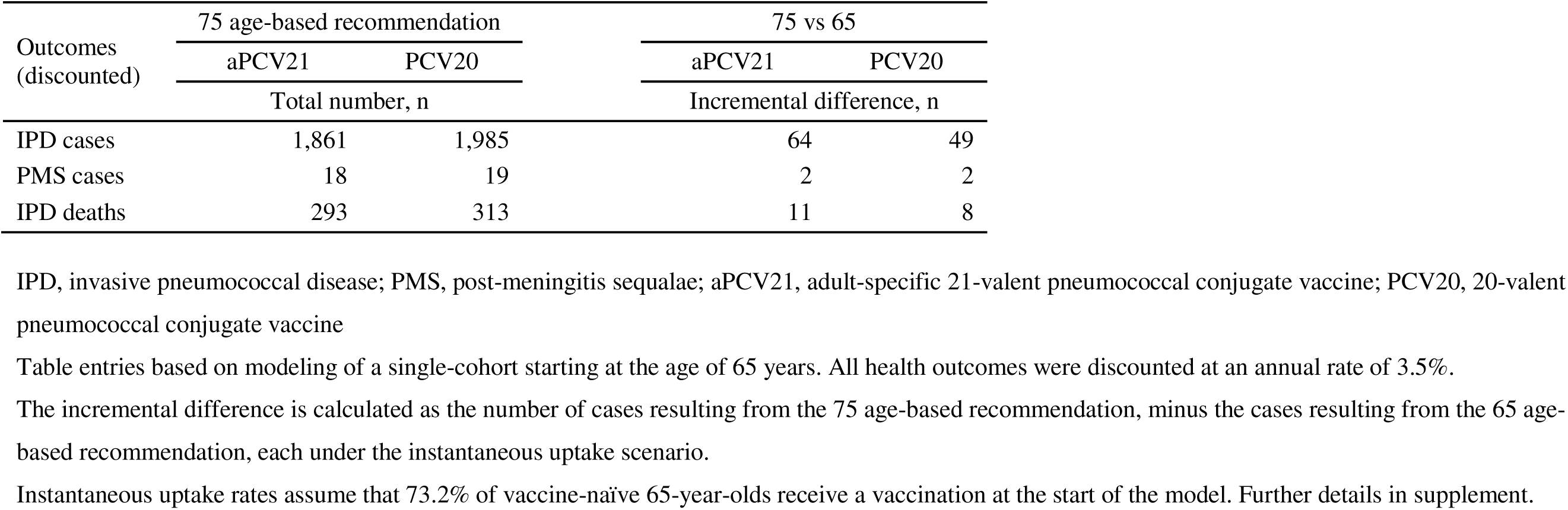
Scenario analysis: Predicted health outcomes at the age of 75 years vs 65 years, using instantaneous vaccination rates, by pneumococcal vaccine type.

Direct medical costs following vaccination with aPCV21, under instantaneous uptake focused solely at 75 years of age, were estimated to be £15.9 million from IPD and £288 thousand from PMS. Direct medical costs following vaccination with PCV20, under instantaneous uptake focused solely at 75 years of age, were estimated to be £16.9 million from IPD and £301 thousand from PMS. Under the age-based recommendation of 75 years and vaccination with aPCV21 (or PCV20, respectively), the impact of shifting from the real-world to instantaneous uptake rate assumptions yielded a 4.1% (2.4%) relative decrease in PMS medical costs and a 2.3% (1.5%) relative decrease in IPD medical costs (**Figure 1**).

### Scenario analysis results: Maximal vaccine impact and optimal vaccination age

In a single-cohort of adults aged 65 years using real-world vaccine uptake rates, the largest reduction in IPD cases occurred with a vaccination age of 65 for both aPCV21 (474 IPD cases averted) and PCV20 (325 IPD cases averted; **Supplemental Figure S2a**). When the standard discounting was removed, the largest reduction in IPD cases occurred with a vaccination age of 75 for both aPCV21 (702 IPD cases averted) and PCV20 (409 IPD cases averted; **Supplemental Figure S2b**).

### Sensitivity analysis results

The DSA indicated that the finding of cost savings for aPCV21 compared with PCV20 was consistent for each input range assumption, as the incremental direct medical costs remained below £ -900 million across all variations of input parameter ranges (**Figure 2**). For all parameters except vaccine efficacy, the predicted incremental cost of using aPCV21 instead of PCV20 ranged from approximately -£1.33 to -£1.08 million. Vaccine efficacy against non-ST3 IPD among low-risk and at-risk adults had the greatest influence on incremental costs. When vaccine efficacy against non-ST3 IPD among at-risk adults was lowered from the base case value of 75.0% to 41.0%, incremental costs went from -£1.20 million to -£0.92 million; whereas, when vaccine efficacy was set to 91.0% incremental costs expanded to -£1.33 million. A similar pattern was observed for efficacy against non-ST3 IPD among high-risk adults.

**Figure 2.**
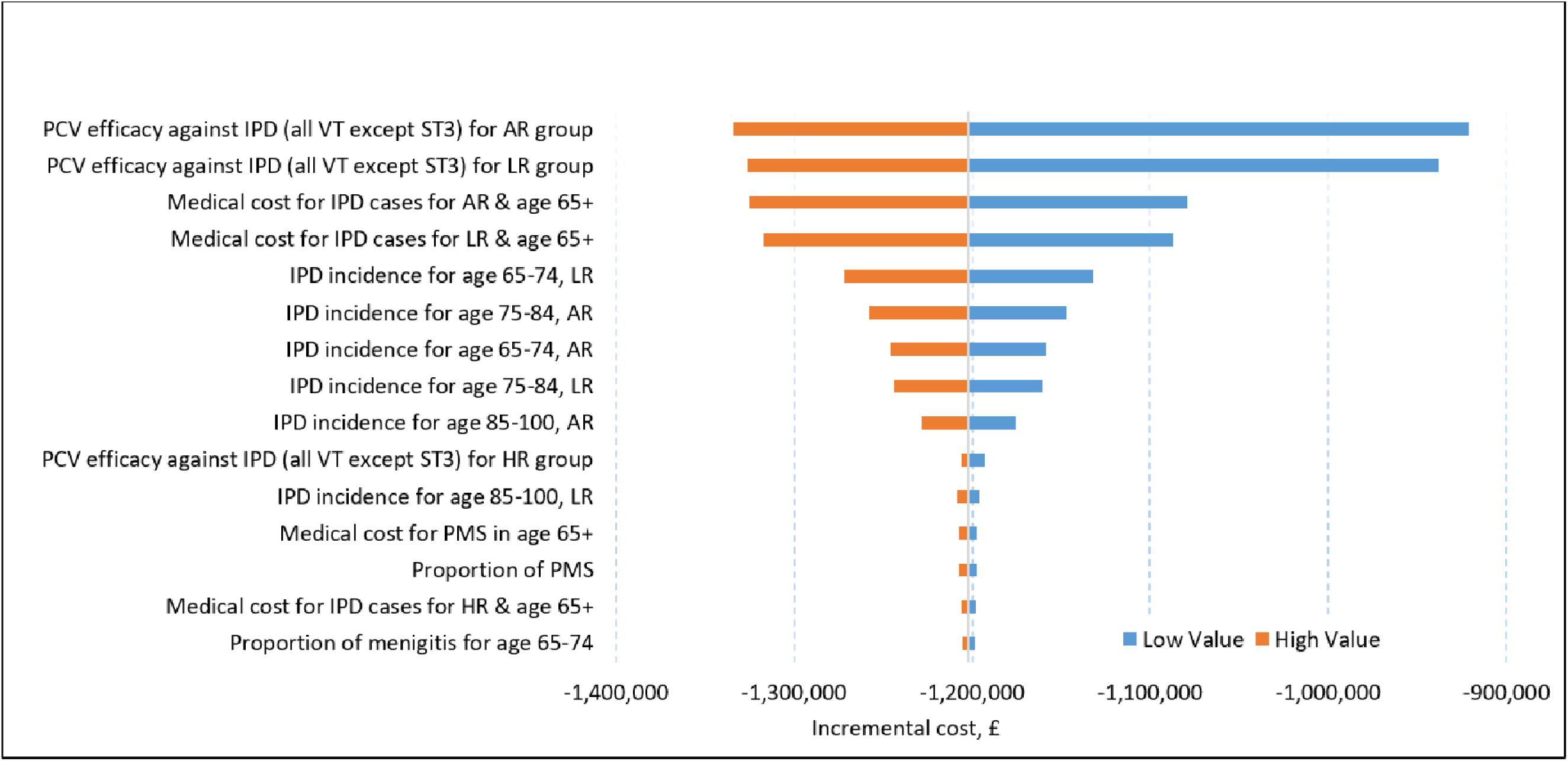
One-way deterministic sensitivity analysis: Top 15 inputs with the greatest influence on the incremental cost for aPCV21 compared with PCV20 with age-based recommendations starting at age 65 years. AR, at-risk; HR, high-risk; IPD, invasive pneumococcal disease; LR, low-risk; PCV, pneumococcal conjugate vaccine; ST3, serotype 3; VT, vaccine type; aPCV21, adult-specific 21-valent pneumococcal conjugate vaccine; PCV20, 20-valent pneumococcal conjugate vaccine. The horizontal axis indicates the incremental, which is the difference in the total direct medical costs under aPCV21 vaccination vs PCV20 under the real-world uptake rate assumption. In the base case scenario, the incremental cost was predicted to be -£1.20 million.

## Discussion

We used a published Markov model to evaluate the health and economic impacts of vaccinating older adults in the UK with aPCV21 or PCV20. Our findings show that aPCV21 averted more IPD cases, PMS cases, IPD deaths, and direct medical costs than PCV20. Specifically, if age-based vaccination starts at 65 years, aPCV21 would avert 138 more IPD cases, 1 more PMS case, and 22 more IPD deaths than PCV20. Our model also predicted that changing the age-based recommendations for adult vaccination from 65 to 75 years would lead to worse discounted health and economic outcomes, independent of the vaccine choice.

Under the age-based recommendation of 65 years, aPCV21 led to fewer IPD cases, PMS cases, and IPD deaths than PCV20. This finding aligns with those of other models evaluating PD outcomes in older adults in Germany, Japan, the UK, and the US.^5,17,25–27^ These other models also showed that the greater impact of aPCV21 on the burden of PD extended to NBPP outcomes.^26,27^ Notably, aPCV21 aligns well with the serotypes that cause adult PD. UK surveillance data show that aPCV21 has higher coverage than PCV20 for IPD serotypes among adults aged ≥65 years (aPCV21 vs PCV20, 84.05% vs 59.65%).^14^ Other data have shown that aPCV21’s unique serotypes account for a substantial proportion of IPD in adults aged ≥50 years in multiple countries.^28–34^ In addition, data from a trial of older adults show that aPCV21 elicited immunological responses to a broader range of pneumococcal serotypes than PPSV23 or PCV15, suggesting the greater potential of aPCV21 to prevent disease.^35^

Our analysis showed that keeping the age-based recommendation at 65 years, instead of 75 years, would avert more IPD cases and deaths for both vaccines, but particularly for aPCV21. In our analysis, a larger incremental difference, across age-based recommendations, in IPD cases was predicted for aPCV21 than for PCV20 (120 vs 92). This is consistent with serotype coverage patterns, which skew toward older age groups more for aPCV21 than for PCV20. As shown in a study by Nikhab et al.,^14^ compared with PCV20, aPCV21 covers 29.1% more IPD cases than PCV20 among adults aged 65-74 years, 43.5% more cases among adults aged 75-84 years, and 54.3% more cases among adults aged 85 years or older.

Few studies have quantitatively assessed the effect of age at vaccination on the burden of PD in older adults. A modeling study of PCVs and PPSV23 by Thindwa et al. reported that the optimal age of pneumococcal vaccination in UK adults is 70 years,^16^ while a study by Chen et al. found that vaccination at age 70 years, instead of 65 years in Australian adults led to fewer IPD deaths.^36^ Only after removing both the discounting of IPD cases and applying the instantaneous uptake rate assumption in our model, did we find that 75 was the optimal age of vaccination (rather than 65). The number of cases averted in Thindwa et al. was higher than what was predicted in the current study, likely caused by the higher IPD incidence input value they used. However, neither Thindwa et al. nor Chen et al. compared the impacts of age-based recommendations on health outcomes under more gradual or “real-world” vaccine uptake rates or with the recommended health outcome discounting.

A more direct comparison of our results on age-based recommendations can be made with a model developed by Danelian et al.^37^ Using model inputs based on 2018-2019 surveillance data from the UK, Danelian et al. found that vaccinating at age 75 averted slightly more IPD cases than vaccinating at age 65. Their model had similar assumptions as ours regarding the waning of vaccine efficacy, which we assumed reached 0% after only 15 years. Notably, with a more gradual waning of vaccine efficacy, the benefit of vaccination becomes more correlated with remaining life-expectancy. Thus, vaccinating later in life is less detrimental under our assumption of waning efficacy than under an assumption of a more gradual decrease, such as that used by Thindwa et al.^16^ This difference in the waning function may explain why Thindwa et al. identified 70 as the optimal age of vaccination, whereas our undiscounted model and Danelian et al.’s model (which only discounted costs and QALYs) identified 75 as the optimal age.

Aligning with our cost findings for aPCV21, several modeling studies have reported that a switch from PCV20 to aPCV21 would result in cost savings for adult vaccination.^25,27,38,39^ US studies have reported that switching from PCV20 to aPCV21 for adults aged 65 years would be economically favorable,^38^ leading to direct medical cost savings of $269 million.^25^ Similarly, a model of older Japanese adults found that switching to aPCV21 would save ¥733 million in direct medical costs over a lifetime horizon.^27^ A model from the Netherlands showed that aPCV21 is cost-effective and would lead to substantial cost savings compared with PCV20.^39^ As with our study, other cost models of aPCV21 assumed that adults were vaccine-naïve and that serotype-specific vaccine efficacy was equivalent or similar for aPCV21 and PCV20,^25,27,38,39^ in accordance with clinical trial findings.^13^ Nevertheless, model methodologies varied, including indirect protection from childhood vaccination, vaccine costs, population demographics, and the use of a societal versus a payer perspective. Thus, our findings are not directly comparable with those of other models.

The current study has limitations that should be considered. Some model assumptions may have simplified the complex dynamics of PD. Our model was static and did not account for dynamic factors, such as herd immunity, serotype replacement, and changes in the force of infection. However, most modeling studies assessing cost-effectiveness and public health impacts of vaccines use a static Markov approach and thus share this limitation. In addition, our results are consistent with those of a more complex dynamic transmission model study.^5^ The risk-based vaccine uptake rates spanning ages 65-74 years were assumed to be constant and equivalent to those for individuals aged 2-64 years, which may not be the case. The incremental differences between aPCV21 and PCV20 may be greater than those predicted in the current study, as serotype distribution data used to inform the model included coverage of 15C by both aPCV21 and PCV20; however, PCV20 does not directly protect against serotype 15C. Another limitation is that our study did not normalize the results by the total number of vaccinations given by the two age-based recommendation cutoffs. Instead, we used uptake rates based on a percentage of the age group being vaccinated. Additionally, the lack of a more gradual or natural waning in vaccine efficacy (as seen in Thindwa et al.^16^) may have predicted an optimal vaccination age that may be older than the true optimum. Lastly, our model did not assume a direct correlation between age and the immune response to the vaccine; doing so would almost certainly make the 75 age-based recommendation less beneficial.

The current study also had important strengths, including using a model that incorporated variation in vaccine efficacy by risk status and age-specific values for vaccine uptake. Furthermore, model inputs for IPD incidence and mortality were derived from surveillance data representative of the UK population. Other model inputs were derived from published reports, which are reliable and transparent data sources. Lastly, the variety of scenario analyses performed suggested the variations across models and assumptions therein are less impactful to the optimal age of vaccination than the decision of whether to discount health outcomes.

In summary, findings of our Markov model indicate that using aPCV21 instead of PCV20 to vaccinate older adults in the UK would prevent more IPD cases and deaths and lead to substantial savings in direct medical costs for IPD. Increasing the age of vaccination from 65 years to 75 years was found to be unfavorable for both health and economic impacts. These findings align with those of other modeling studies of aPCV21, highlighting the potential role of this vaccine for the UK’s adult pneumococcal vaccination program.

## Data Availability

All data produced in the present work are contained in the manuscript

## Acknowledgments

The authors thank Ingrid Peterson, PhD, in collaboration with ScribCo for medical writing assistance.

## Funding

This study was funded by Merck Sharp & Dohme LLC, a subsidiary of Merck & Co., Inc., Rahway, NJ, USA.

## Supplementary Material

**Supplementary section:** Risk distribution methodology

To make our model more accurate, a recent improvement to the risk progression methodology was made to our model. A separate demographic model was fit to risk distribution data to estimate annual risk progression rates between the low-risk, at-risk, and high-risk groups. Using a combination of risk distribution data from van Hoek et al. 2012^20^ and the 2024-2025 PPV coverage report, the risk distribution for ages 2-64 was 91.6% low-risk, 6.8% at-risk, and 1.7% high-risk, and the risk distribution for ages 65+ was 47.8% low-risk, 47.7% at-risk, and 4.6% high-risk. The at-risk group included asplenia/splenic dysfunction, chronic respiratory disease, chronic heart disease, chronic kidney disease, chronic liver disease, and diabetes; the high-risk group included Immunosuppression (excluding asplenia), cochlear implants, and cerebrospinal fluid leaks.

The demographic model consisted of 3 risk progression pathways: one from low-risk to at-risk, another from low-risk to high-risk, and a final option from at-risk to high-risk. Each of these pathways had a progression probability as a function of time (t) with coefficient (A) and an annual percent change (r):

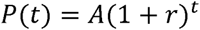

Using demographic data from **Supplemental Table S1**, this demographic model was simulated in “Microsoft® Excel® for Microsoft 365 MSO (Version 2604 Build 16.0.19929.20172) 64-bit” with annual time steps.

Using the Solver Add-in, the GRG nonlinear solving method was used to minimize the sum of squared errors between the risk distribution data in **Table 1** (also reported at the start of this section) and the calculated risk distribution values from the demographic model. The coefficients of the risk transition probabilities from low-risk to at-risk, low-risk to high-risk, and at-risk to high-risk were found to be 0.0312%, 0.0100%, and 0.2263%, respectively; the annual percent change were found to be 6.913%, 0.000%, and 2.597%, respectively. For convenience of replication of this model or for others to use the results of this demographic model, we have included the annual transition probabilities and the resultant risk distributions by age in **Supplemental Table S5**.

**Supplementary section:** Vaccine uptake rates methodology

Real-world uptake rates were taken directly from the 2024-2025 PPV coverage report published by UK Health Security Agency (UKHSA).^24^ The values used are reported in the first column of Supplemental Table S2. We assumed the uptake rates, which are provided based on the recent vaccination rates across different age cohorts, would be a reasonable approximation for the future year-by-year uptake rates for the simulated cohort of 65-year-olds. This is a limitation of the model but appears reasonable based on the relatively stable uptake rates over the past several PPV coverage reports. Using historic rates of a single-aged-cohort was one option to have consistent rates, but these might be less reflective of what is to be expected for future uptake rates.

An additional set of assumptions was that the uptake rates beyond 75 years-of-age were fixed at a constant rate of 1.8% and that no further uptake was simulated beyond age 90 under the 65 age-based recommendations. This enabled relatively consistent overall coverage between the age-based recommendations as this allowed the 1.8% uptake rate to spawn ages 85 through 99 in the 75 age-based recommendation scenario.

The alternative set of vaccine uptake scenarios assumed a focused uptake rate at the start of the age-based recommendation. A resultant uptake rate of 73.2% (occurring at either 65 years-of-age or 75, based on whether we were simulating the 65 or 75 age-based recommendation) was calculated using the same 2024-2025 PPV coverage report, by first subtracting out the non-vaccine-naïve population (137,928 of the 65-year-old cohort that were vaccinated prior to 1 April 2024) from both the total number of people registered and the total number of people vaccinated on or before 31 March 2025 – yielding 73.2% (8,251,388/11,264,962).

---

**Supplemental Table S1.** Population size and all-cause mortality by age.

| Age, years | Population size | Mortality |
| --- | --- | --- |
| 65 | 689,198 | 0.009850 |
| 66 | 687,334 | 0.010541 |
| 67 | 674,764 | 0.011629 |
| 68 | 651,687 | 0.012729 |
| 69 | 652,398 | 0.013960 |
| 70 | 660,817 | 0.015134 |
| 71 | 672,642 | 0.016573 |
| 72 | 702,415 | 0.017603 |
| 73 | 753,009 | 0.020780 |
| 74 | 575,023 | 0.023002 |
| 75 | 549,939 | 0.025161 |
| 76 | 540,477 | 0.028006 |
| 77 | 494,925 | 0.031290 |
| 78 | 435,050 | 0.036236 |
| 79 | 383,368 | 0.040204 |
| 80 | 386,890 | 0.044888 |
| 81 | 373,288 | 0.049594 |
| 82 | 350,944 | 0.055479 |
| 83 | 322,102 | 0.062849 |
| 84 | 292,999 | 0.070876 |
| 85 | 264,997 | 0.080151 |
| 86 | 231,822 | 0.090342 |
| 87 | 206,428 | 0.100117 |
| 88 | 185,189 | 0.113679 |
| 89 | 161,430 | 0.127235 |
| 90 | 128,003 | 0.139064 |
| 91 | 110,202 | 0.155719 |
| 92 | 93,042 | 0.170281 |
| 93 | 77,199 | 0.187869 |
| 94 | 62,695 | 0.208374 |
| 95 | 41,995 | 0.230457 |
| 96 | 32,317 | 0.251479 |
| 97 | 24,190 | 0.272484 |
| 98 | 17,599 | 0.303821 |
| 99 | 12,252 | 0.311631 |
| ≥100 | 15,539 | 1.000000 |

Source for population estimates: UK Office of National Statistics.<sup>18</sup> Source for mortality data: UK Office of National Statistics.<sup>19</sup>

**Supplemental Table S2.** Real-world pneumococcal vaccine uptake rates by age and age-based vaccination recommendations.

| Vaccine uptake rate (%) | Age at which the single-cohort of 65-year-olds receives the listed uptake rate |  |
| --- | --- | --- |
|  | 65 age-based recommendation | 75 age-based recommendation<sup>A</sup> |
| 15.4 | 65 | 75 |
| 12.4 | 66 | 76 |
| 6.9 | 67 | 77 |
| 5.5 | 68 | 78 |
| 4.6 | 69 | 79 |
| 5.2 | 70 | 80 |
| 4.2 | 71 | 81 |
| 3.0 | 72 | 82 |
| 2.5 | 73 | 83 |
| 2.4 | 74 | 84 |
| 1.8 | 75-89 | 85-99 |

Vaccine uptake rates by age were based on published data for the existing age-based recommendation.<sup>24</sup>
<sup>A</sup> Risk-based vaccination was assumed to occur between the ages of 65-74 years for this strategy.

**Supplemental Table S3.**
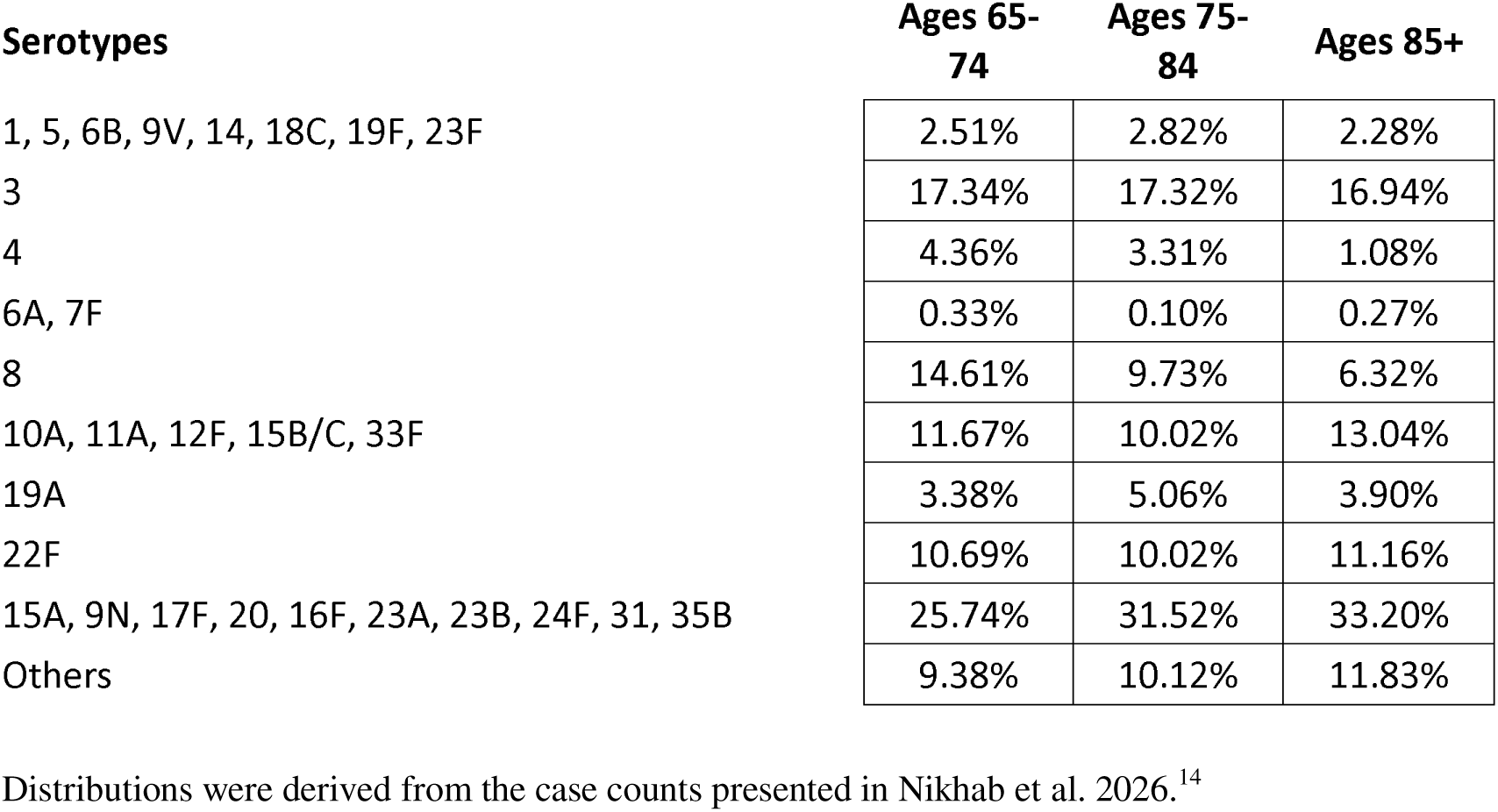
The distribution of pneumococcal serotypes in cases of invasive pneumococcal disease for people aged ≥65 years.

| **Serotypes** | **Ages 65-74** | **Ages 75-84** | **Ages 85+** |
| --- | --- | --- | --- |
| 1, 5, 6B, 9V, 14, 18C, 19F, 23F | 2.51% | 2.82% | 2.28% |
| 3 | 17.34% | 17.32% | 16.94% |
| 4 | 4.36% | 3.31% | 1.08% |
| 6A, 7F | 0.33% | 0.10% | 0.27% |
| 8 | 14.61% | 9.73% | 6.32% |
| 10A, 11A, 12F, 15B/C, 33F | 11.67% | 10.02% | 13.04% |
| 19A | 3.38% | 5.06% | 3.90% |
| 22F | 10.69% | 10.02% | 11.16% |
| 15A, 9N, 17F, 20, 16F, 23A, 23B, 24F, 31, 35B | 25.74% | 31.52% | 33.20% |
| Others | 9.38% | 10.12% | 11.83% |

Distributions were derived from the case counts presented in Nikhab et al. 2026.<sup>14</sup>

**Supplemental Table S4.** Effect ranges for parameters assessed in the one-way deterministic sensitivity analysis.

| Inputs | Description | Base Case | Lower Bound | Upper Bound |
| --- | --- | --- | --- | --- |
| Vaccine efficacy (PCVs) | Against IPD (all VT except ST3) for LR group | 0.750 | 0.414 | 0.908 |
|  | Against IPD (all VT except ST3) for AR group | 0.750 | 0.414 | 0.908 |
|  | Against IPD (all VT except ST3) for HR group | 0.250 | 0.138 | 0.303 |
|  | Against IPD due to ST3 for LR group | 0.260 | 0.000 | 0.534 |
|  | Against IPD due to ST3 for AR group | 0.260 | 0.000 | 0.534 |
|  | Against IPD due to ST3 for HR group | 0.087 | 0.000 | 0.178 |
| IPD incidence (cases per 100,000 person-years) | Age 65-74 | 13.65 | 10.92 | 16.38 |
|  | Age 75-84 | 24.89 | 19.91 | 29.87 |
|  | Age ≥85 | 44.69 | 35.75 | 53.62 |
| Proportion of meningitis | Age 65-74 | 0.03 | 0.03 | 0.04 |
|  | Age 75-84 | 0.02 | 0.02 | 0.03 |
|  | Age ≥85 | 0.01 | 0.01 | 0.01 |
| Proportion of PMS | Proportion of PMS out of IPD meningitis | 0.44 | 0.36 | 0.53 |
| IPD fatality rate | Age ≥65 | 0.157 | 0.126 | 0.189 |
| Direct Treatment Cost for IPD (2024 GBP) | Age ≥65 | 8544.61 | 6835.69 | 10253.53 |
| Direct Treatment Cost for PMS (2024 GBP) | Age ≥65 | 1554.80 | 1243.84 | 1865.76 |
| Discount rate (per year) | Discount rate for health outcomes | 0.035 | 0.00 | N/A<sup>A</sup> |
|  | Discount rate for costs | 0.035 | 0.00 | 0.05 |

AR, average-risk; HR, high-risk; IPD, invasive pneumococcal disease; LR, low-risk; PCV, pneumococcal conjugate vaccine; PMS, post-meningitis sequelae
<sup>A</sup> The discount rate for health outcomes was not used in the one-way deterministic sensitivity analysis, as it does not impact the costs. A value of 0 was used as part of the scenarios.

**Supplemental Table S5.** Derived age-specific risk-distributions and transition probabilities.

| Age | Risk distribution |  |  | Risk transition probabilities |  |  |
| --- | --- | --- | --- | --- | --- | --- |
|  | Low-risk | At-risk | High-risk | Low-risk to at-risk | Low-risk to high-risk | At-risk to high-risk |
| 65 | 0.690 | 0.278 | 0.033 | 0.0241 | 0.0005 | 0.0023 |
| 66 | 0.673 | 0.294 | 0.033 | 0.0258 | 0.0005 | 0.0023 |
| 67 | 0.655 | 0.310 | 0.035 | 0.0275 | 0.0006 | 0.0023 |
| 68 | 0.637 | 0.328 | 0.036 | 0.0294 | 0.0006 | 0.0023 |
| 69 | 0.618 | 0.346 | 0.037 | 0.0315 | 0.0006 | 0.0023 |
| 70 | 0.598 | 0.364 | 0.038 | 0.0336 | 0.0006 | 0.0023 |
| 71 | 0.577 | 0.384 | 0.039 | 0.0360 | 0.0006 | 0.0023 |
| 72 | 0.556 | 0.403 | 0.040 | 0.0385 | 0.0006 | 0.0023 |
| 73 | 0.535 | 0.424 | 0.042 | 0.0411 | 0.0006 | 0.0023 |
| 74 | 0.512 | 0.445 | 0.043 | 0.0440 | 0.0007 | 0.0023 |
| 75 | 0.489 | 0.466 | 0.044 | 0.0470 | 0.0007 | 0.0023 |
| 76 | 0.466 | 0.488 | 0.046 | 0.0502 | 0.0007 | 0.0023 |
| 77 | 0.442 | 0.511 | 0.047 | 0.0537 | 0.0007 | 0.0023 |
| 78 | 0.418 | 0.533 | 0.048 | 0.0574 | 0.0007 | 0.0023 |
| 79 | 0.394 | 0.556 | 0.050 | 0.0614 | 0.0008 | 0.0023 |
| 80 | 0.369 | 0.579 | 0.052 | 0.0657 | 0.0008 | 0.0023 |
| 81 | 0.345 | 0.602 | 0.053 | 0.0702 | 0.0008 | 0.0023 |
| 82 | 0.320 | 0.625 | 0.055 | 0.0750 | 0.0008 | 0.0023 |
| 83 | 0.296 | 0.647 | 0.056 | 0.0802 | 0.0008 | 0.0023 |
| 84 | 0.272 | 0.670 | 0.058 | 0.0858 | 0.0009 | 0.0023 |
| 85 | 0.248 | 0.692 | 0.060 | 0.0917 | 0.0009 | 0.0023 |
| 86 | 0.225 | 0.713 | 0.062 | 0.0980 | 0.0009 | 0.0023 |
| 87 | 0.203 | 0.733 | 0.064 | 0.1048 | 0.0009 | 0.0023 |
| 88 | 0.182 | 0.753 | 0.065 | 0.1121 | 0.0010 | 0.0023 |
| 89 | 0.161 | 0.772 | 0.067 | 0.1198 | 0.0010 | 0.0023 |
| 90 | 0.142 | 0.789 | 0.069 | 0.1281 | 0.0010 | 0.0023 |
| 91 | 0.123 | 0.806 | 0.071 | 0.1370 | 0.0010 | 0.0023 |
| 92 | 0.106 | 0.821 | 0.073 | 0.1464 | 0.0011 | 0.0023 |
| 93 | 0.091 | 0.834 | 0.075 | 0.1565 | 0.0011 | 0.0023 |
| 94 | 0.076 | 0.847 | 0.077 | 0.1674 | 0.0011 | 0.0023 |
| 95 | 0.064 | 0.857 | 0.079 | 0.1789 | 0.0011 | 0.0023 |
| 96 | 0.052 | 0.867 | 0.081 | 0.1913 | 0.0012 | 0.0023 |
| 97 | 0.042 | 0.875 | 0.083 | 0.2045 | 0.0012 | 0.0023 |
| 98 | 0.033 | 0.882 | 0.085 | 0.2187 | 0.0012 | 0.0023 |
| 99 | 0.026 | 0.887 | 0.087 | 0.2338 | 0.0013 | 0.0023 |
| 100 | 0.020 | 0.891 | 0.089 | 0.2500 | 0.0013 | 0.0023 |

**Supplemental Figure S1.**
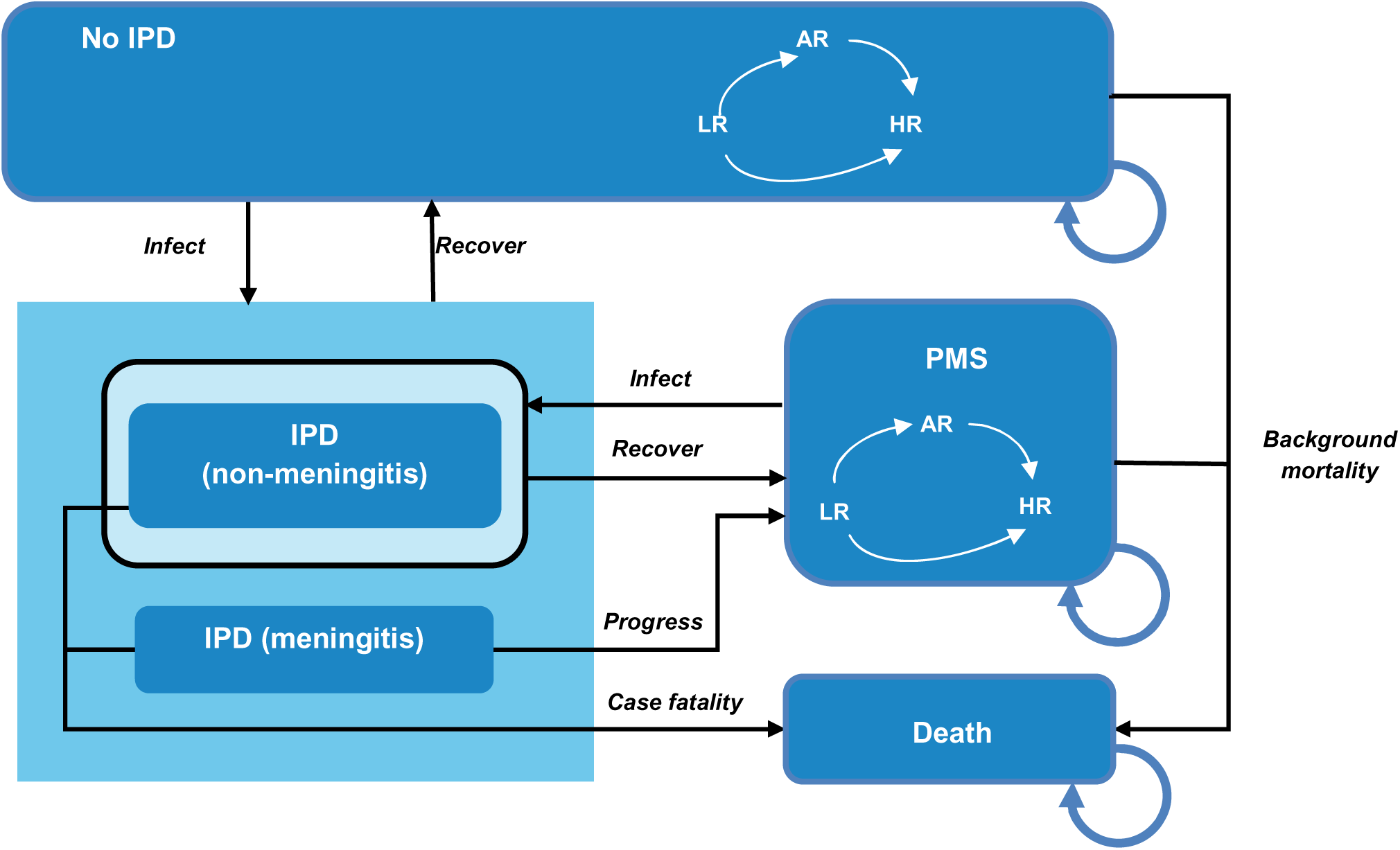
Structure of the Markov model. AR, at-risk; HR, high-risk; IPD, invasive pneumococcal disease; LR, low-risk; PMS, post-meningitis sequelae Figure adapted from Yi et al (2024).^17^

**Supplemental Figure S2.**
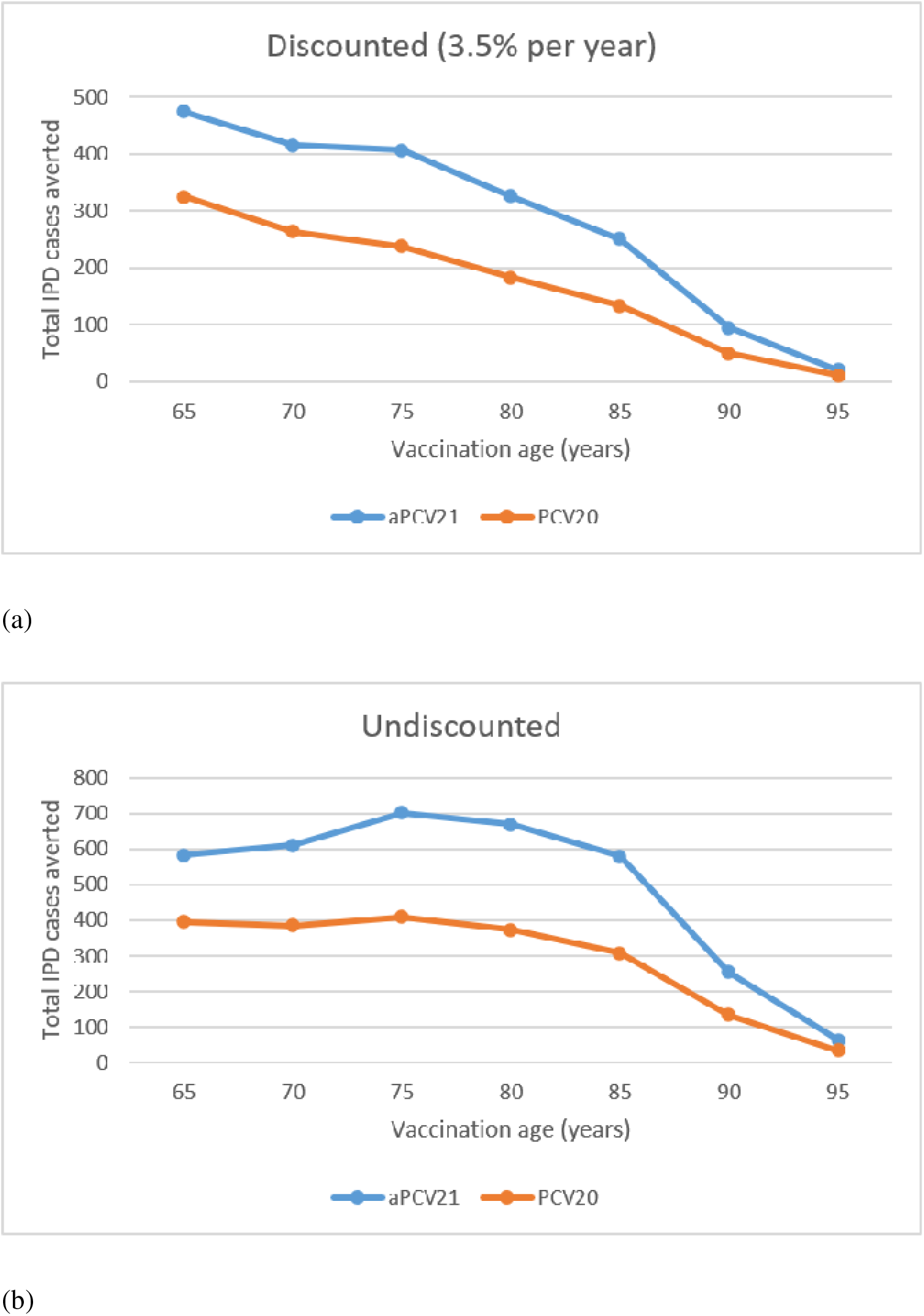
Scenario analysis results for maximal vaccine impact and optimal vaccination age: Total lifetime IPD cases averted in a single-cohort of adults aged 65 year, by vaccination age. IPD, invasive pneumococcal disease; aPCV21, adult-specific 21-valent pneumococcal conjugate vaccine; PCV20, 20-valent pneumococcal conjugate vaccine A cohort of 689,198 vaccine-naïve adults aged 65 years were simulated until a maximum age of 100. The total cases averted was defined as the difference between no vaccination coverage and 100% vaccination coverage at the specified vaccination ages. Results are presented under both (a) standard discounting rates (all health outcomes discounted at 3.5% per year) and (b) without discounting outcomes.

## Notes

### Competing Interest Statement

All authors are employees of Merck Sharp & Dohme LLC, a subsidiary of
Merck & Co., Inc., Rahway, NJ, USA and MSD (UK) Limited, London, UK.
aPCV21 was developed by Merck & Co., Inc., Rahway, NJ, USA.

